# Overground robotic walker use in the home and community: a six-month prospective cohort study

**DOI:** 10.1101/2025.06.07.25329084

**Authors:** Alicia J Hilderley, Christa M Diot, Hua Shen, Sean P Dukelow, Kelly A Larkin-Kaiser, Adam Kirton, Elizabeth G Condliffe

## Abstract

**Importance:** Outcomes following long-term use of overground robotic walkers have not been studied, even though children with mobility impairments are using these devices for extended periods.

**Objective:** To evaluate the impacts of six months of overground robotic walker use in the home and community.

**Design:** Observational cohort study.

**Setting:** Home and community.

**Participants:** Volunteer sample of participants with mobility impairments who privately obtained an overground robotic walker. 255 device users were contacted, 171 consented to participate.

**Exposure:** Six months of participant-initiated overground robotic walker use.

**Main outcomes and measures:** The primary outcome of functional ability was assessed by parent report using the Gillette Functional Assessment Questionnaire (FAQ). Secondary outcome measures were parent-reported physical activity, positive affect, sleep disturbance, and bowel movement frequency. Questionnaires were sent digitally at baseline (when users received device use training), and then 1-Month, 3-Months and 6-Months later. The device tracked monthly usage, specifically number of steps, minutes of use, average cadence (steps/minute), and the number of times the device was used.

**Results:** Median participant age was 6 years (range 1 to 24), 42.1% were female, 70.8% had a diagnosis of cerebral palsy, and most were not independently ambulatory (97.3% of participants who reported function). Adjusted cumulative link mixed models demonstrated a significant main effect of time for FAQ scores, with increased log odds of a higher FAQ score at each time point (ß=0.86, 95% CI [0.25, 1.46], *p*=0.006). Adjusted repeated measures linear mixed-effects models demonstrated significant main effects of time for secondary outcomes, with improvements in physical activity scores (ß=0.96, 95% CI [0.21, 1.71], p=0.012), sleep disturbance scores (ß=-0.82, 95% CI [-1.61, -0.04], *p*=0.040), average cadence (steps/minute) (ß=1.86, 95% CI [0.61, 3.11], *p*=0.004), and also decreases in the number of times the device was used per month (ß=-0.95, 95% CI [-1.63, -0.26], *p*=0.007). Device usage time and total steps per month did not significantly change over time.

**Conclusions and Relevance:** Six months of overground robotic walker use resulted in significant improvements in functional ability and secondary outcomes linked to physical inactivity. Device usage time was consistent over time, suggesting feasibility of long-term home and community use.

## INTRODUCTION

Severe mobility impairments limit opportunities for physical activity and increase the risk of sequelae associated with sedentary behaviours.^1,2^ Standard physical interventions have questionable effectiveness,^3^ highlighting the need for new approaches such as robot-assisted devices. In fact, robot-assisted gait training devices may be more effective than standard interventions for improving gait-specific outcomes.^4,5^ However this evidence is largely based on devices that are tethered to a treadmill system, upon which a user walks with facilitated leg movement and body-weight support. These devices typically limit use to clinical environments and require trained clinicians to operate.

Overground robot-assisted walking devices may address limitations of tethered devices. These devices are suitable for use on flat surfaces within the home and community. The opportunity for family-initiated use has been introduced by an overground robotic walker commercially available to families. Devices are designed for children with mobility impairments and have robotic legs with an external power source attached to a stable four wheeled walker. Families can use and control the device independently, removing reliance on a therapist and clinical space for upright mobility. This may increase feasibility of greater frequency and dose,^6^ but patterns of overground robot-assisted walking device use beyond six weeks have not been reported.^7,8^

However, families are not waiting for evidence of device effectiveness, as proven by the lease or purchase of over 500 devices from one company. Research involving the use of overground robotic walking devices is growing, with indications of potential benefits^7,9^ and at least three clinical trials underway (NCT05473676; NCT05463211; NCT05378243). Potential benefits of overground robotic walker use may extend beyond common foci of gait and functional ability to address secondary complications of limited mobility including physical inactivity, low mood, poor sleep, and bowel function problems.^7^ At this time, characterizing users, patterns of real-world home and community device use, and identifying who may benefit most from long-term use is necessary to inform and justify the momentum behind this rehabilitation technology.

We conducted a cohort study to determine if six months of real-world overground robotic walker use could improve functional ability and secondary complications of limited mobility.

## METHOD

This was an observational prospective single cohort study. The ethics committee of the University of Calgary (Calgary, Canada) approved the protocol. Informed consent/assent was electronically obtained from participants/guardians. This study followed the Strengthening the Reporting of Observational Studies in Epidemiology (STROBE) reporting guideline for cohort studies.^10^

### Participants

A volunteer sample of participants who had committed to a minimum 6-month lease or purchase of a Trexo device from the Trexo Robotics company (Mississauga, Canada) were recruited if they: (i) had a reliable internet connection for data collection; and (ii) signed our research study electronic informed consent form. There were no criteria regarding participant age, diagnosis, medical history, or country of residence. The number of participants consented during the study timeline from June 2020 to December 2023 determined the sample size.

### Device Use

This observational study did not prescribe device use. Rather, the amount of Trexo use was dictated by the participants and their families. The Trexo can be used indoors and outdoors on flat and stable surfaces, therefore participants and families could choose to use the device in their home, school and/or community.

Real-world data from each participant’s Trexo device were collected and stored on a tablet associated with a unique serial number per device. Once connected to Wi-Fi, data were automatically uploaded to Amazon QuickSight (Amazon, Seattle, USA). Each time the device was used, the tablet recorded: usage time when walking, number of steps taken, and average cadence (steps per minute). Monthly averages for each variable were calculated using R Statistical Software (v4.2.2; R Core Team 2021) based on a 28-day epoch for consistency.

### Parent-Reported Outcome Measures

This study was initiated during the COVID-19 pandemic and therefore used remote data collection methods, which also reduced participant burden and eliminated geographical restrictions. All outcome measures were collected electronically using Jotform (Jotform, Vancouver, Canada) or Qualtrics (Qualtrics, Provo, USA) and completed by participants’ parents or guardians. Parent-reported outcome measures were used to ensure consistency in the role of respondent. Outcomes were assessed at four time points. The baseline assessment was completed close to the time users received their device and users received training on home use from the Trexo Robotics company. The subsequent three assessments were completed one month, three months, and six months after the training date. Email and SMS reminders were sent to parents/guardians to increase response rates. If an assessment was not completed, parents were still invited to complete subsequent assessments.

Demographic data collected at baseline included participant age, sex, primary diagnosis, Gross Motor Function Classification System (GMFCS) Level, functional mobility using the Functional Mobility Scale (FMS), and spasticity/tone. Parent-reported diagnoses were categorized as cerebral palsy, genetic condition, acquired brain injury, spinal cord injury, or other. The GMFCS Family Report Questionnaire was used to classify children and youth with physical disabilities into five functional categories ranging from I (walks without limitations) to V (transported by others in a manual wheelchair).^11^ The FMS was used to describe functional mobility over three distinct distances (5, 50, and 500 meters), representing home, school, and community settings. Each distance has scores of 1 (uses wheelchair) to 6 (independent on all surfaces) with additional categories C (crawling) and N (does not complete this distance). Parents were asked to report any medication or dietary strategy use (yes or no) related to bowel function and whether medications were used to manage spasticity/tone (yes or no).

Outcome measures were collected at all four assessments. The primary outcome was functional ability assessed by the Gillette Functional Assessment Questionnaire: Functional Walking Scale (FAQ). The FAQ is a 10-level parent-report of walking ability. A score of 1 indicates no stepping ability and 10 indicates independent walking and climbing on varied terrain without difficulty.^12^ Minimal important difference for the FAQ is 1.5 points.^13^ Sleep disturbance, positive affect, and physical activity were evaluated using Parent Proxy Item Banks (v1.0) of the Patient-Reported Outcome Measurement Information System (PROMIS® National Institutes of Health, Bethesda, MD, USA). Specific item banks used were the Sleep Disturbance—Short Form 8a^14^, Positive Affect—Short Form 8a^15^, and Physical Activity—Short Form 4a^16^. Questions were based on a seven-day recall period. Scores were converted to a standardized T-score with mean 50 (SD 10). T-scores >50 indicate above the mean for positive affect and physical activity, but below the mean (worse) for sleep disturbance. Finally, parents reported their child’s bowel function over the previous week. Bowel movement frequency had five options: more than daily; daily; every other day; two times in the last week; and less than two in the last week.

### Statistical Analysis

Participant demographics were summarized using the appropriate descriptive statistics. To model changes over time and identify baseline predictors, repeated measures linear mixed-effects models were fit for continuous outcomes and cumulative link mixed models were fit for ordinal outcomes. In the cumulative link mixed models, the estimated coefficients represent the log odds (i.e. the logarithm of the odds ratio) of being in a higher ordinal category for each one unit increase in the predictor. In the repeated measures linear mixed effects models, estimated coefficients represent the average outcome change for each one unit increase in the predictor. Models included a random intercept for participants. Separate unadjusted and adjusted models were run for each outcome of interest.

Unadjusted models considered the fixed effect of time. Adjusted models considered fixed effects of time, age, sex, diagnosis of cerebral palsy, progressive condition, and baseline FAQ score ≤2. To identify baseline predictors of change over time, interaction effects of time with other fixed effects were quantified. For these interaction effects, a monthly estimate was calculated by summing the estimate of the interaction effect with the estimate of the main effect of time. The significance for all tests was set at *p*<0.05. All available data were included without imputation for missing values.

Statistical analysis was completed in R (version 4.3.3).^17^

## RESULTS

From June 11, 2020, to December 8, 2023, a total of 171 participants were enrolled out of the 255 device users contacted. The median age of participants was 6 years (range 1 to 24 years), and 42.1% were female. The majority of participants had a diagnosis of cerebral palsy (70.8%). Twenty-one participants (12.3%) had a progressive condition (e.g. spinal muscular atrophy, Rett syndrome). Most participants were primarily wheelchair users: 145 of 149 of participants who reported GMFCS level were not independently ambulatory (Level IV or V) and all participants who completed the FMS 500m used a wheelchair or stroller (FMS 500m = 1 or N). Common FAQ scores were “1”: cannot take any steps (29.8%), or “2”: can do some stepping with assistance (40.9%). High tone was reported for 40.9% of participants, with 36.8% using medication to control spasticity at baseline. Bowel movement frequency was at minimum every other day for 61.1% of participants, with 52.0% using medication or dietary strategies to support their bowel function. The median T-score at baseline for physical activity was 41.40 (range 31.3 to 61.3) and for sleep disturbance was 63.80 (range 43.9 to 84.1), and the mean T-score for positive affect was 47.83 (SD 9.16). Further participant characteristics are presented in Table 1.

**Table 1.** Baseline Participant Characteristics. GMFCS: Gross motor function classification system; FMS: Functional mobility scale, FAQ: Gillette Functional Assessment Questionnaire: Functional Walking Scale.

| **Outcome** | **Baseline values** |
| --- | --- |
| Age (years) | Median 6 (IQR 6), range 1 to 24 |
| Gender (n) | 99 Male (57.9%)<br>72 Female (42.1%) |
| Primary diagnosis (n) | 121 Cerebral palsy (70.8%)<br>36 Genetic condition (21.21%)<br>3 Acquired brain injury (1.8%)<br>2 Spinal cord injury (1.2%)<br>9 Other/unknown (5.3%) |
| Progressive (n) | 150 Non progressive (87.7%)<br>21 Progressive (12.3%) |
| GMFCS Level (n) | 106 Level V (62%)<br>39 Level IV (22.8%)<br>3 Level III (1.8%)<br>1 Level II (0.6%)<br>22 Not reported (12.9%) |
| FMS 5m (n) | 104 Does not complete (60.8%)<br>14 Crawls (8.2%)<br>12 Uses wheelchair (7.0%)<br>3 Uses walker or frame (1.8%)<br>38 Not reported (22.2%) |
| FMS 50m (n) | 113 Does not complete (66.1%)<br>14 Uses wheelchair (8.2%)<br>3 Uses walker or frame (1.8%)<br>41 Not reported (24.0%) |
| FMS 500m (n) | 129 Does not complete (75.4%)<br>8 Uses wheelchair (4.7%)<br>34 Not reported (19.9%) |
| FAQ (n) | 51 Score 1 (29.8%)<br>70 Score 2 (40.9%)<br>13 Score 3 (7.6%)<br>1 Score 4 (0.6%)<br>2 Score 5 (1.2%)<br>1 Score 6 (0.6%)<br>1 Score 7 (0.6%)<br>32 Not reported (18.7%) |
| Positive affect (T-score) | Mean 47.83 (SD 9.16), range 21.60 to 67.30 |
| Sleep disturbance (T-score) | Median 63.80 (IQR 10.1), range 43.90 to 84.10 |
| Physical activity (T-score) | Median 41.40 (IQR 9.70), range 31.30 to 61.30 |
| Bowel movement frequency (n) | 5 Once a week (2.9%)<br>18 Twice a week (10.5%)<br>39 Every other day (22.8%)<br>64 Every day (37.4%)<br>17 More than daily (9.9%)<br>28 Not reported (16.4%) |
| Bowel movement dietary strategy/medication use (n) | 55 No (32.2%)<br>89 Yes (52.0%)<br>27 Not reported (15.8%) |
| Spasticity medication use (n) | 73 No (42.7%)<br>63 Yes (36.8%)<br>25 Not reported (20.5%) |

Of the 171 participants, 169 completed at least one questionnaire timepoint and/or had device usage data for at least one month. Baseline questionnaires were completed by 144 (84.21%) participants (median 7.5 [IQR 21.0] days before Trexo training). 107 (62.57%) participants completed the Month 1 questionnaires (median 36.0 [IQR 16.0] days after training), 95 (55.55%) participants completed the Month 3 questionnaires (median 101.0 [IQR 21.5] days after training), and 99 (57.89%) participants completed the Month 6 questionnaires (median 197 [IQR 33.0] days after training). Monthly device usage data was available for maximum 165 participants (Month 1) to minimum 136 participants (Month 6). Table 2 presents monthly medians for outcome measures and device usage variables. Median participant use of their robotic walking aids was 118 to 172 minutes/month with 3136 to 5434 steps/month.

**Table 2.**
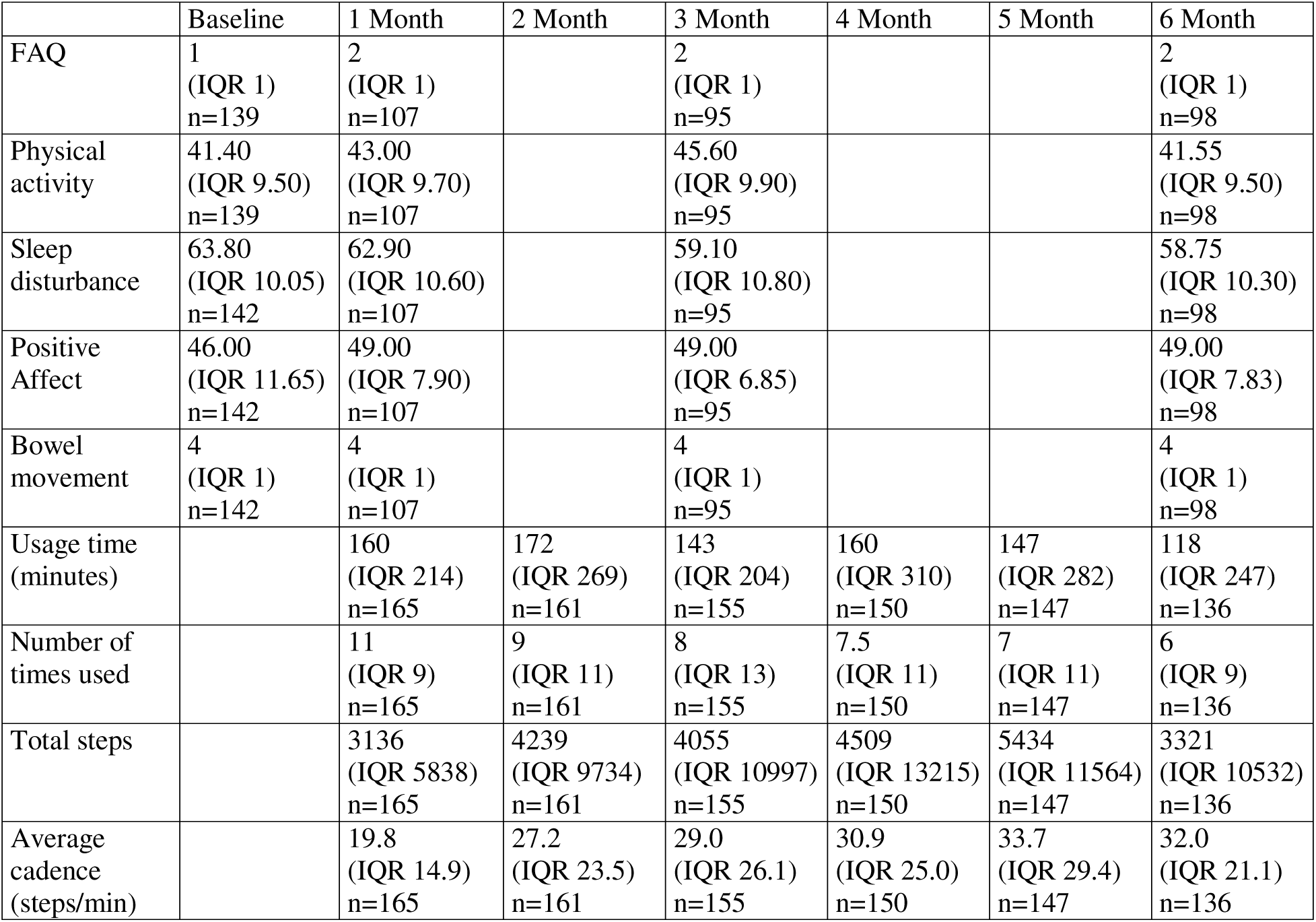
Median Outcome Measure Scores. Median values presented with interquartile ranges (IQR); FAQ: Gillette Functional Assessment Questionnaire: Functional Walking Scale.

Results of the unadjusted model indicated a significant main effect of time on the primary outcome of functional ability. Each additional month increased the log odds of a higher FAQ score by 0.14 (95% CI [0.14, 0.15], *p*<0.001). After adjusting for age, sex, diagnosis, progressive condition, baseline function, and their interactions with time, the main effect of time remained significant and was strengthened (ß=0.86, 95% CI [0.25, 1.46], *p*=0.006). See Figure 1 and Table 3.

**Figure 1.**
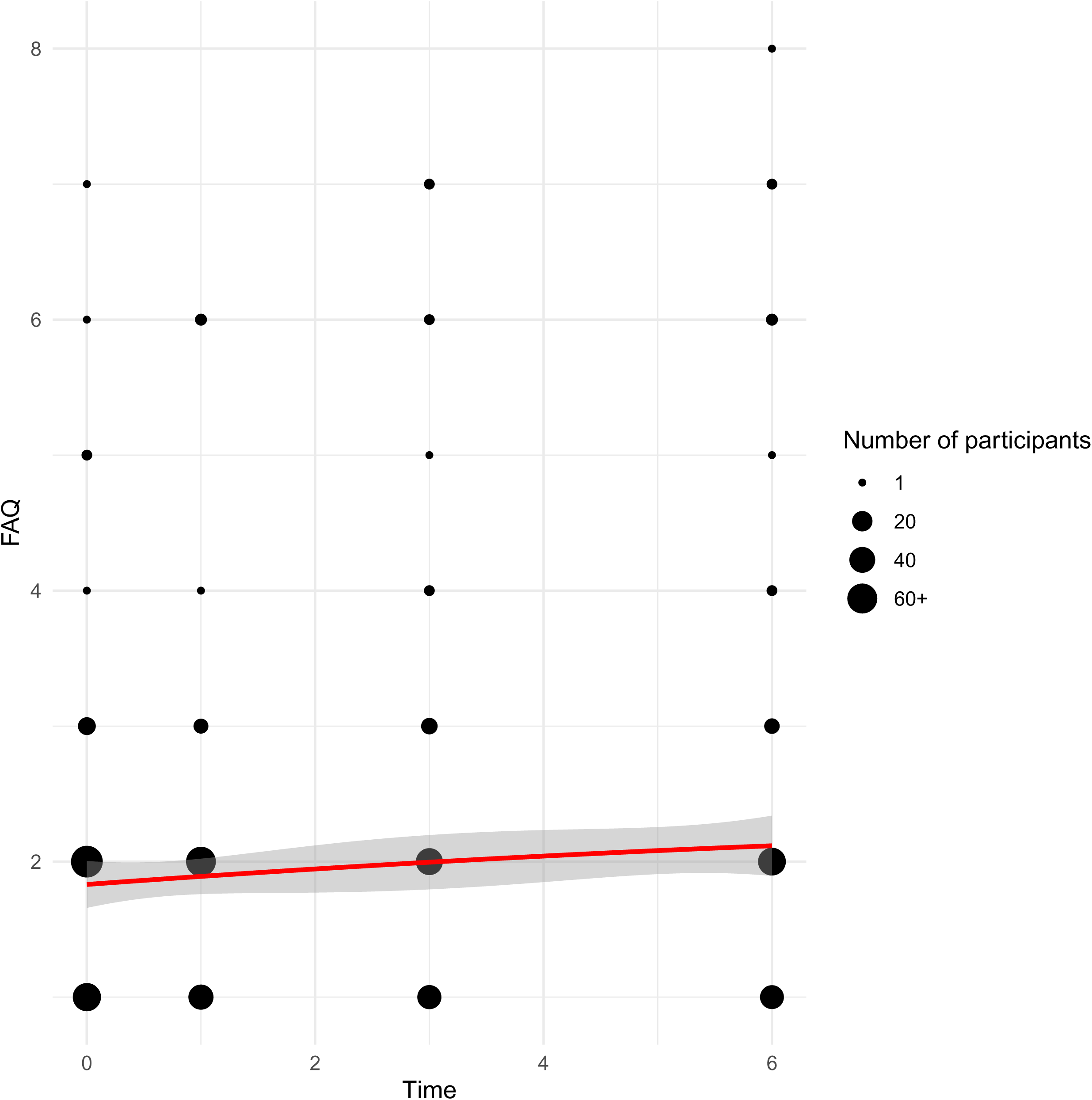
Changes over Time in Functional Ability. Changes in the Gillette Functional Assessment Questionnaire (FAQ) scores over time, presented in months. Dot size is continuous and scaled proportionally to the actual number of participants.

**Table 3.** Estimated Effects of Time on Outcomes from Unadjusted and Adjusted Models. Bolded text highlights significant values; estimates are log odds for ordinal data, units for continuous data; FAQ: Gillette Functional Assessment Questionnaire: Functional Walking Scale.

| Dependent | Unadjusted model<br>ß (95% CI) | *p*-value | Adjusted model<br>ß (95% CI) | *p*-value |
| --- | --- | --- | --- | --- |
| FAQ | **0.14 (0.14, 0.15)** | **<0.001** | **0.86 (0.25, 1.46)** | **0.006** |
| Physical Activity | **0.29 (0.04, 0.53)** | **0.024** | **0.96 (0.21, 1.71)** | **0.012** |
| Positive Affect | **0.43 (0.21, 0.65)** | **<0.001** | 0.33 (-0.34, 1.01) | 0.333 |
| Sleep Disturbance | **-0.57 (-0.77, -0.36)** | **<0.001** | **-0.82(-1.61, -0.04)** | **0.040** |
| Bowel movements | **0.20 (0.10, 0.29)** | **<0.001** | 0.24 (-0.15, 0.62) | 0.227 |
| Usage Time | **-6.73(-12.71, -0.75)** | **0.028** | -9.97 (-30.01, 10.06) | 0.327 |
| Times Used | **-0.94 (11.72, 14.71)** | **<0.001** | **-0.95 (-1.63, -0.26)** | **0.007** |
| Total Steps | **410.89 (104.20, 717.58)** | **0.009** | 161.00 (-722.37, 1044.38) | 0.721 |
| Average Cadence | **2.03 (19.75, 25.35)** | **<0.001** | **1.86 (0.61, 3.11)** | **0.004** |

For unadjusted models of secondary outcomes over time, significant improvements were observed over time in all secondary outcomes (see Table 3). Changes in physical activity (ß=0.96, 95% CI [0.21, 1.71], *p*=0.012) and sleep disturbance (ß=-0.82, 95% CI [-1.61, -0.04], *p*=0.040) remained significant after adjustment.

In the unadjusted models of device usage variables, significant increases were observed in total monthly steps and average cadence, while significant decreases were observed in usage time and number of times used. After adjustment, changes over time in times used (ß=-0.95, 95% CI [-1.63, - 0.26], *p*=0.007) and average cadence (ß=1.86, 95% CI [0.61, 3.11], *p*=0.004) remained significant.

Baseline predictors of change over time in clinical outcomes and device use were identified by significant interactions between time and predictors, when adjusting for all other predictors. See Table 4 for values and monthly estimates. Participants with a baseline FAQ score of ≤2 improved less on the FAQ over time as compared to participants with a baseline FAQ>2 (log odds ß= -0.44, 95% CI [-0.86, -0.02], *p*=0.042). There was a negative relationship between age and changes over time in FAQ score (log odds ß= -0.05, 95% CI [-0.09, -0.01], *p*=0.022), meaning older participants improved less on FAQ scores. Males had greater improvements in FAQ scores over time compared to females (log odds ß=0.46, 95% CI [0.11, 0.82], *p*=0.011) and had greater decreases in sleep disturbance scores (ß=-0.49, 95% CI [-0.93, -0.06], *p*=0.027). Participants with cerebral palsy had smaller reductions in sleep disturbance scores as compared to participants with other diagnoses (ß=0.74, 95% CI [0.19,1.29], *p*=0.009), indicating less improvements in sleep. Participants with a progressive condition had slower increases in bowel movement frequency over time (log odds ß=-0.40, 95% CI [-0.72, -0.07], *p*=0.018) and less decreases in sleep disturbance scores (ß=0.69, 95% CI [0.01,1.37], *p*=0.047) as compared to participants with non-progressive conditions. The were no significant predictors of device use over time.

**Table 4.** Interaction Effects Between Baseline Predictors and Time and on Changes in Outcome Measures. Negative interaction effect estimates indicate a decrease over time, while positive interaction effect estimates indicate an increase over time; bolded text highlights significant values; estimates are log odds for ordinal data, units for continuous data; FAQ: Gillette Functional Assessment Questionnaire: Functional Walking Scale; CP: cerebral palsy.

| Baseline Predictor<br>(Interaction with Time) | Outcome Measure | Interaction Effect $\beta$<br>(95% CI) | *p*-value | Interaction effect $\beta$<br>per time point<br>(95% CI) |
| --- | --- | --- | --- | --- |
| Baseline FAQ $\leq 2$ | **FAQ** | **-0.44 (-0.86, -0.02)** | **0.042** | 0.42 (-0.09, 0.94) |
|  | Physical activity | -0.31 (-1.03, 0.40) | 0.388 | 0.65 (-0.30, 1.59) |
|  | Positive affect | -0.02 (-0.66, 0.62) | 0.944 | 0.31 (-0.54, 1.16) |
|  | Sleep disturbance | -0.43 (-1.02, 0.16) | 0.158 | -1.25 (-1.87, -0.63) |
|  | Bowel movement frequency | 0.10 (-0.18, 0.39) | 0.474 | 0.34 (0.39, 0.64) |
| Age | **FAQ** | **-0.05 (-0.09, -0.01)** | **0.022** | 0.56 (0.04, 1.08) |
|  | Physical activity | -0.06 (-0.12, 0.00) | 0.051 | 0.60 (-0.05, 1.25) |
|  | Positive affect | -0.02 (-0.07, 0.04) | 0.495 | 0.30 (-0.36, 0.80) |
|  | Sleep disturbance | 0.04 (-0.01, 0.09) | 0.093 | -0.57 (-1.27, 0.14) |
|  | Bowel movement frequency | 0.01 (-0.02, 0.03) | 0.445 | 0.29 (-0.05, 0.63) |
| Male sex | **FAQ** | **0.46 (0.11, 0.82)** | **0.011** | 1.32 (0.60, 2.03) |
|  | Physical activity | 0.40 (-0.13, 0.92) | 0.137 | 1.36 (0.58, 2.14) |
|  | Positive affect | 0.27 (-0.21, 0.74) | 0.270 | 0.60 (-0.10, 1.30) |
|  | **Sleep disturbance** | **-0.49 (-0.93, -0.06)** | **0.027** | -1.32 (-2.19, -0.44) |
|  | Bowel movement frequency | -0.09 (-0.30, 0.11) | 0.377 | 0.14 (-0.29, 0.57) |
| CP Diagnosis | FAQ | -0.36 (-0.80, 0.08) | 0.110 | 0.50 (0.03, 0.96) |
|  | Physical activity | -0.56 (-1.22, 0.11) | 0.101 | 0.41 (-0.21, 1.02) |
|  | Positive affect | 0.07 (-0.53, 0.66) | 0.826 | 0.40 (-0.16, 0.96) |
|  | **Sleep disturbance** | **0.74 (0.19, 1.29)** | **0.009** | -0.08 (-0.75, 0.58) |
|  | Bowel movement frequency | -0.12 (-0.38, 0.15) | 0.395 | 0.12 (-0.20, 0.44) |
| Progressive condition | FAQ | 0.16 (-0.38, 0.70) | 0.567 | 1.01 (0.36, 1.67) |
|  | Physical activity | -0.46 (-1.27, 0.36) | 0.274 | 0.50 (-0.26, 1.27) |
|  | Positive affect | 0.10 (-0.628, 0.84) | 0.780 | 0.44 (-0.25, 1.12) |
|  | **Sleep disturbance** | **0.69 (0.01, 1.37)** | **0.047** | -0.14 (-0.96, 0.68) |
|  | **Bowel movement frequency** | **-0.40 (-0.72, -0.07)** | **0.018** | -0.16 (-0.55, 0.23) |

After adjusting for all other predictors, baseline participant characteristics were identified that significantly predicted clinical outcome scores at all time points. Participants with lower baseline function, assessed by FAQ≤2, had lower FAQ scores at all time points (log odds ß=-5.09, 95% CI [- 7.67, -2.51], *p<*0.001) and higher physical activity scores at all time points (ß=4.26, 95% CI [0.78, 7.75], *p*= 0.018). The were no other significant baseline predictors of clinical outcomes or device use. See eTable 1 for results of all fixed and interaction effects.

## DISCUSSION

We found that six months of overground robotic walker use significantly improved functional ability, physical activity, and sleep disturbance in a group of participants who were primarily wheelchair users.

Our results indicate that improvements in functional ability are possible outside of clinical settings throughout childhood and adolescence. Significant improvements in functional ability following targeted interventions are rarely achieved for children with limited ability to walk.^18^ For children with cerebral palsy, who were the largest population in this study at 70.8%, gross motor function is typically stable after seven years of age.^19^ Here we observed improvements in a cohort aged one to 21 years old, with greater functional gains over time for younger ages. It is important to note that the gains observed in the current study were modest and statistical significance does not indicate that gains will be experienced by all participants. However, it is promising that on average, users were expected to improve their functional ability following six-months of family-directed overground robotic walker use in the home and community.

To identify participants who may benefit most from robot-assisted walking, we evaluated baseline predictors of changes over time in functional ability. Baseline FAQ score, age, and sex were significant predictors of FAQ changes over time. The lack of significant relationship with CP diagnosis highlights the potential for children with significant mobility limitations due to other causes to improve functional abilities with device use. Interestingly, there were no significant relationships between having a progressive condition and change in FAQ score. Individuals with progressive conditions are often excluded from controlled trials, which is not justified by our results.

The importance of significant improvements in secondary outcomes must also be underscored, as impairments in sleep, affect, physical activity, and bowel function are reported to negatively affect quality of life.^20–22^ Before adjusting for baseline predictors, all secondary outcomes significantly improved over time. Following adjustment, only improvements over time in physical activity and sleep disturbance remained significant. Improving sleep is highly meaningful for the perceived well-being of children and of parents.^23^ Understanding that baseline characteristics influence changes over time in positive affect and bowel movements may be useful for decision-making when these outcomes are a therapeutic target.

Many assistive technologies have high non-adoption and abandonment rates,^24^ challenges which were not reflected in device usage results. Usage time and total steps per month did not significantly change over time in the adjusted models, suggesting successful integration of device use into family routines over six months. Increased ease of device use may have contributed to the significant improvements observed in average cadence and the decrease in monthly times used. There were no baseline predictors of device use, indicating equal potential for device use across baseline age, diagnoses, and functional abilities.

Considering the high cost of devices, family decisions and clinician recommendations to obtain devices need to be carefully weighed. Randomized controlled trials are underway to provide the necessary level of evidence to inform clinical purchase,^25^ which will importantly support equitable access. Examination of supports required for robotic walker use and economic analyses will be key study components to inform access expansion to all children who may benefit from robot-assisted walking.

### Limitations

This study reports the largest sample size in pediatric robot-assisted walking research, yet a main limitation of the study is the missing data. We chose to analyze all collected data using mixed-effects models to accommodate missing data and avoid waste. In our analysis, individuals with incomplete observations at some time points still contribute partial information for the time points where data are available, thereby allowing the mixed effects model to incorporate all usable data without discarding participants entirely for having missing measurements. Missing data related to usage could arise from a technical problem with data upload, Wi-Fi connection issues, or no device usage – reasons which cannot be distinguished from the data. The sample with device usage data decreased from n=165 at Month 1 to n=136 at Month 6, but whether this is due to data upload issues or no device use is unknown. No device use could have occurred for child and family-related reasons, like surgery, shifting priorities, or dislike of the device. External factors may also have contributed. For example, in seasons with extreme heat or cold, participants using the device outdoors may have ceased use. Reduced access to community spaces, delays in fitting required equipment such like ankle-foot-orthoses, and supply chain issues for required equipment may have impacted use during the pandemic.

Potential bias was introduced by the observational design, enrolling families who privately leased or purchased the devices and therefore may have been predisposed to observing improvements. While using the GMFCS across diagnoses is common, it is a classification system specific to cerebral palsy.^26^ Variability in cerebral palsy diagnoses may have led to misclassification of some participants.^27^ Finally, indications of improvements in functional ability are promising but for many participants the magnitude of change over this six month study was less than the minimum important change in FAQ score of 1.5.^13^

## CONCLUSIONS

Findings from this prospective observation study suggest that six-months of overground robotic walker use can result in improvements in functional ability and secondary outcomes linked to physical inactivity. Children who are primarily wheelchair users can benefit from long-term home and community use of robot-assisted walking devices.

## Data Availability

All data produced in the present study are available upon reasonable request to the authors.

## Author Contributions

EGC was the principal investigator of the study and conceived the trial. EGC, SPD, KALK, CD contributed to the study design. AJH and CD enrolled participants. AJH and CD collected the data. AJH and HS analyzed data. AJH, HS, and EGC interpreted the data. AJH and HS accessed and verified the data. AJH and ECG wrote the manuscript. All authors contributed to the revision of the manuscript. All authors vouch for the fidelity of the protocol and the accuracy and completeness of the reported data. All authors reviewed and approved of the manuscript before submission. AJH, HS, and EGC had full access to all the data in the study and take responsibility for the integrity of the data and the accuracy of the data analysis. We thank the children and families for dedicating time and effort to participate in this study.

## References

1. Jonsson U, Eek MN, Sunnerhagen KS, Himmelmann K. Health Conditions in Adults With Cerebral Palsy: The Association With CP Subtype and Severity of Impairments. Front Neurol. 2021;12:732939.

2. Dum R, Walter V, Thomas NJ, Krawiec C. Odds of Cardiometabolic Diseases and Medications in Children With Cerebral Palsy. J Child Neurol. 2023;38(3–4):239–46.

3. Ryan JM, Cassidy EE, Noorduyn SG, O’Connell NE. Exercise interventions for cerebral palsy. Cochrane Database Syst Rev. 2017;6(6):CD011660.

4. Conner BC, Remec NM, Lerner ZF. Is robotic gait training effective for individuals with cerebral palsy? A systematic review and meta-analysis of randomized controlled trials. Clin Rehabil. 2022;2692155221087084.

5. Cortés-Pérez I, González-González N, Peinado-Rubia AB, Nieto-Escamez FA, Obrero-Gaitán E, García-López H. Efficacy of Robot-Assisted Gait Therapy Compared to Conventional Therapy or Treadmill Training in Children with Cerebral Palsy: A Systematic Review with Meta-Analysis. Sensors (Basel). 2022;22(24):9910.

6. van Hedel HJA, Meyer-Heim A. Robot-Assisted Gait Training for Children and Youth with Cerebral Palsy. In: Miller F, Bachrach S, Lennon N, O’Neil ME, editors. Cerebral Palsy. Springer International Publishing; 2020. p. 2797–816.

7. Diot CM, Youngblood JL, Friesen AH, Wong T, Santos TA, Norman BM, et al. Robot-Assisted Gait Training with Trexo Home: Users, Usage and Initial Impacts. Children (Basel). 2023;10(3):437.

8. Choi JY, Kim SK, Hong J, Park H, Yang S seung, Park D, et al. Overground Gait Training With a Wearable Robot in Children With Cerebral Palsy: A Randomized Clinical Trial. JAMA Network Open. 2024;7(7):e2422625.

9. Diot CM, Thomas RL, Raess L, Wrightson JG, Condliffe EG. Robotic lower extremity exoskeleton use in a non-ambulatory child with cerebral palsy: a case study. Disabil Rehabil Assist Technol. 2021;1–5.

10. von Elm E, Altman DG, Egger M, Pocock SJ, Gøtzsche PC, Vandenbroucke JP. Strengthening the reporting of observational studies in epidemiology (STROBE) statement: guidelines for reporting observational studies. BMJ. 2007;335(7624):806–8.

11. Morris C, Galuppi BE, Rosenbaum PL. Reliability of family report for the Gross Motor Function Classification System. Developmental Medicine & Child Neurology. 2004;46(7):455–60.

12. Novacheck TF, Stout JL, Tervo R. Reliability and validity of the Gillette Functional Assessment Questionnaire as an outcome measure in children with walking disabilities. J Pediatr Orthop. 2000;20(1):75–81.

13. Ammann-Reiffer C, Bastiaenen CHG, Van Hedel HJA. Measuring change in gait performance of children with motor disorders: assessing the Functional Mobility Scale and the Gillette Functional Assessment Questionnaire walking scale. Dev Med Child Neurol. 2019;61(6):717–24.

14. Forrest CB, Meltzer LJ, Marcus CL, de la Motte A, Kratchman A, Buysse DJ, et al. Development and validation of the PROMIS Pediatric Sleep Disturbance and Sleep-Related Impairment item banks. Sleep. 2018;41(6).

15. Forrest CB, Ravens-Sieberer U, Devine J, Becker BD, Teneralli R, Moon J, et al. Development and Evaluation of the PROMIS® Pediatric Positive Affect Item Bank, Child-Report and Parent-Proxy Editions. J Happiness Stud. 2018;19(3):699–718.

16. Tucker CA, Bevans KB, Becker BD, Teneralli R, Forrest CB. Development of the PROMIS Pediatric Physical Activity Item Banks. Phys Ther. 2020;100(8):1393–410.

17. R Core Team. R: A language and environment for statistical computing. Vienna, Austria: R Foundation for Statistical Computing; 2024.

18. Novak I, Morgan C, Fahey M, Finch-Edmondson M, Galea C, Hines A, et al. State of the Evidence Traffic Lights 2019: Systematic Review of Interventions for Preventing and Treating Children with Cerebral Palsy. Curr Neurol Neurosci Rep. 2020;20(2).

19. Hanna SE, Rosenbaum PL, Bartlett DJ, Palisano RJ, Walter SD, Avery L, et al. Stability and decline in gross motor function among children and youth with cerebral palsy aged 2 to 21 years. Dev Med Child Neurol. 2009;51(4):295–302.

20. Horwood L, Li P, Mok E, Oskoui M, Shevell M, Constantin E. Health-related quality of life in Canadian children with cerebral palsy: what role does sleep play? Sleep Medicine. 2019;54:213– 22.

21. Belsey J, Greenfield S, Candy D, Geraint M. Systematic review: impact of constipation on quality of life in adults and children. Aliment Pharmacol Ther. 2010;31(9).

22. Colver A, Rapp M, Eisemann N, Ehlinger V, Thyen U, Dickinson HO, et al. Self-reported quality of life of adolescents with cerebral palsy: a cross-sectional and longitudinal analysis. The Lancet. 2015;385(9969):705–16.

23. Mörelius E, Hemmingsson H. Parents of children with physical disabilities – perceived health in parents related to the child’s sleep problems and need for attention at night. Child: Care, Health and Development. 2014;40(3):412–8.

24. Howard J, Fisher, Zoe, Kemp, Andrew H., Lindsay, Stephen, Tasker, Lorna H., and Tree JJ. Exploring the barriers to using assistive technology for individuals with chronic conditions: a meta-synthesis review. Disabil Rehabil Assist Technol. 2022;17(4):390–408.

25. Calabrò RS, Müller-Eising C, Diliberti ML, Manuli A, Parrinello F, Rao G, et al. Who Will Pay for Robotic Rehabilitation? The Growing Need for a Cost-effectiveness Analysis. Innov Clin Neurosci. 2020;17(10–12):14–6.

26. Towns M, Rosenbaum P, Palisano R, Wright FV. Should the Gross Motor Function Classification System be used for children who do not have cerebral palsy? Dev Med Child Neurol. 2018;60(2):147–54.

27. Aravamuthan BR, Fehlings DL, Novak I, Gross P, Alyasiry N, Tilton AH, et al. Uncertainties Regarding Cerebral Palsy Diagnosis: Opportunities to Clarify the Consensus Definition. Neurol Clin Pract. 2024;14(6):e200353.

