## Supplemental Material for "Overground robotic walker use in the home and community: a six-month prospective cohort study"

This supplement has been provided by the authors to give readers additional information about the work.

### eTable 1. Estimated Effects on Outcomes

| Predictors | FAQ | Physical activity | Positive affect | Sleep disturbance | Bowel movement frequency | Average cadence | Steps | Usage time | Times Used |
|---|---|---|---|---|---|---|---|---|---|
| Time | **0.86 (0.25, 1.46)**** | **0.96 (0.21, 1.71)*** | 0.33 (-0.34, 1.01) | **-0.82 (-1.61, -0.04)*** | 0.24 (-0.15, 0.62) | **1.86 (0.61, 3.11)**** | 161.00 (-722.37, 1044.38) | -9.97 (-30.01, 10.06) | **-0.95 (-1.63, -0.26)**** |
| Age | 0.17 (-0.04, 0.38) | -0.11 (-0.38, 0.17) | 0.170 (-0.16, 0.50) | -0.14 (-0.45, 0.17) | -0.11 (-0.24, 0.02) | -0.42 (-1.06, 0.23) | -268.06 (-795.86, 259.74) | 0.62 (-10.70, 11.94) | -0.01 (-0.36, 0.34) |
| Sex Male | -0.79 (-2.62, 1.04) | 0.62 (-1.77, 3.00) | 1.51 (-1.42, 4.43) | -0.92 (-3.65, 1.80) | 0.94 (-0.22, 2.10) | 1.21 (-4.58, 7.00) | 1249.65 (-3477.17, 5976.46) | -12.30 (-113.32, 88.72) | -0.41 (-3.51, 2.70) |
| Baseline FAQ ≤2 | **-5.09 (-7.67, -2.51)** *** | **4.26 (0.78, 7.75)*** | 0.67 (-3.61, 4.95) | 2.21 (-1.77, 6.20) | -1.53 (-3.24, 0.18) | 3.83 (-4.51, 12.16) | 3605.13 (-3207.99, 10418.25) | 75.73 (-71.88, 223.34) | 4.32 (-0.16, 8.79) |
| Progressive condition | -0.68 (-3.76, 2.40) | 1.11 (-2.86, 5.09) | 2.66 (-2.25, 7.56) | -3.84 (-8.38, 0.71) | 1.67 (-0.27, 3.61) | 3.11 (-6.45, 12.67) | -264.56 (-8112.64 ,7583.52) | -55.43 (-222.27, 111.40) | 0.06 (-5.07, 5.19) |
| Diagnosis of CP | 1.23 (-1.04, 3.50) | 0.76 (-2.20, 3.72) | 1.22 (-2.43, 4.86) | -2.37 (-5.75, 1.01) | -0.25 (-1.67, 1.17) | 1.19 (-5.98, 8.36) | -1855.13 (-7701.02, 3990.76) | -93.49 (-219.93, 32.95) | -1.99 (-5.83, 1.84) |
| Time: Age | **-0.05 (-0.09, -0.01)*** | -0.06 (-0.12, 0.00) | -0.02 (-0.07, 0.04) | 0.04 (-0.01, 0.09) | 0.01 (-0.02, 0.03) | 0.07 (-0.04, 0.17) | 7.95 (-64.99, 80.88) | -1.24 (-2.87, 0.40) | -0.02 (-0.08, 0.03) |
| Time: Sex Male | **0.46 (0.11, 0.82)*** | 0.40 (-0.13, 0.92) | 0.27 (-0.21, 0.74) | **-0.49 (-0.93, -0.06)*** | -0.09 (-0.30, 0.11) | 0.34 (-0.56, 1.24) | 313.20 (-324.86, 951.26) | 13.76 (-0.49, 28.01) | 0.34 (-0.16, 0.83) |
| Time: FAQ ≤2 Baseline | **-0.44 (-0.86, -0.02)*** | -0.31 (-1.03, 0.40) | -0.02 (-0.66, 0.62) | -0.43 (-1.02, 0.16) | 0.10 (-0.18, 0.39) | 1.24 (-0.05, 2.52) | 194.75 (-711.33, 1100.83) | -1.54 (-22.17, 19.09) | -0.31 (-1.02, 0.39) |
| Time: Progressive | 0.16 (-0.38, 0.70) | -0.46 (-1.27, 0.36) | 0.10 (-0.63, 0.84) | **0.69 (0.01, 1.37)*** | **-0.40 (-0.72, -0.07)*** | -0.40 (-1.88, 1.08) | 379.10 (-664.38, 1422.58) | 15.99 (-7.60, 39.58) | 0.13 (-0.68, 0.94) |
| Time: CP Diagnosis | -0.36 (-0.80, 0.08) | -0.56 (-1.22, 0.11) | 0.07 (-0.53, 0.66) | **0.74 (0.19, 1.29)**** | -0.12 (-0.38, 0.15) | -0.87 (-1.99, 0.25) | -88.96 (-881.74, 703.83) | 9.29 (-8.66, 27.24) | 0.01 (-0.60, 0.63) |

Legend: ß (95% CI) presented; Bolded text highlights significant values; *** $p<0.001$ ** $p<0.01$ * $p<0.05$; estimates are log odds for ordinal data, units for continuous data; FAQ: Gillette Functional Assessment Questionnaire: Functional Walking Scale; Progressive: progressive condition; CP: cerebral palsy.
